# Non-differential risk of SARS-CoV-2 infection for members of polling stations on Catalan parliament voting day

**DOI:** 10.1101/2021.05.13.21257143

**Authors:** Manuel Medina-Peralta, Luis Garcia-Eroles, Eduardo Hermosilla, Antonio Fuentes, Leonardo Méndez-Boo, Francesc Fina, Mireia Fàbregas, Ermengol Coma, Yolanda Lejardi, Marc Ramentol, Ismael Peña-López

## Abstract

We aimed to assess the risk of SARS-CoV-2 infection for polling station members during the Catalan elections in February 2021. We compared the incidence 14 days after the elections between a cohort of polling station members (N= 18,304) and a control cohort paired by age, sex and place of residence. A total of 37 COVID-19 cases (0.2%) were confirmed in the members of the polling stations and 43 (0.23%) in the control group (p-value 0.576). Our study suggests that there was no greater risk of infection for the members of the polling stations.

## Introduction

Elections are a centerpiece of the political system of democratic countries. During COVID-19 pandemic, some elections and referendums have been cancelled or postponed worldwide [1]. Several concerns and potential risks of holding elections have been discussed: the risk of increasing the spread of COVID-19 in the community [2]; and the risk of infection for both voters and members of the polling station committees. To date, no studies have analyzed these possible risks regarding polling station members.

On 14th February 2021, Catalonia (Spain) held its elections to the Catalan parliament’with more than 5.3 million people with the right to vote [3]. To mitigate transmission of COVID-19 at polling locations and to protect both voters and members of polling stations, the Catalan Government had adopted some measures including encouraging mail voting; specific time schedules for COVID-19 cases, close contacts and vulnerables; limitation of people inside polling stations; use of facemasks; use of hand sanitizer; social distancing between voters and between voters and members of polling stations; use of personal protective equipment during COVID-19 cases voting time schedule; outdoor voting if possible; and the offer to polling station members to perform a lateral flow test (LFT) some days before the elections [4].

The aim of our study was to analyze the risks of SARS-CoV-2 infection for polling station members, by comparing their incidence of COVID-19 during the 14 days after the elections to the incidence in a control population.

## Methods

We performed a controlled retrospective cohort study to determine if being a member of a polling station during the Catalan elections was associated with an increased risk of transmission of SARS-CoV-2. In Catalonia, members of polling stations are randomly selected from the census and it’s mandatory unless exception applies. We used data from 14th to 28th February 2021. As the incubation period of COVID-19 ranges from 3 to 14 days [5], we performed a subanalysis dividing the study period into two different periods: the three days after the elections and from day 4 to 14 after the elections.

We compared the incidence of COVID-19 in two cohorts: a sample of members of polling station which data available (convenience sample) and a control group paired by age in years, sex, municipality of residence and risk of COVID-19. Risk of COVID-19 was assessed using the classification of the Catalan Government that classifies patients by risk of severity of COVID-19 according to some socio-demographics and clinical features. This classification includes three levels: low, moderate and high risk.

The sample of members of the polling station were provided by the Direcció General de Participació Ciutadana i Processos Electorals of the Catalan Government who received this information directly from each municipality. Data were further linked to the regional central database of reverse transcriptase polymerase chain reaction (RT-PCR) and lateral flow test (LFT) fo rSARS-CoV2 to determine the incidence of COVID-19 in both cohorts.

### Statistical analysis

An “Exact Matching” (1:1 relation) between polling station cohort and eligible control cohort was performed using the variables described above. We compared the incidence of SARS-CoV-2 infection using the Fisher’s exact test. We considered that there were significant differences between both cohorts when the p-value was lower than 0.05 All analyses were conducted using R version 4.0.0.

### Ethical statement

This work was approved by the Clinical Research Ethics Committee of the IDIAPJGol (project code: 21/104-PCV), including a waiver for the informed consent of citizens taking part in the study.

## Results

During the voting day on 14th February 2021, more than 9,000 polling station committees were constituted that included 27,417 members. Out of them, we finally included 18,304 people (66.8%) from 6,320 different polling station committees.

Both cohorts had a mean age of 41.8 (12.2) years old and were 48.8% female. Regarding the risk of COVID-19, 59.7% had low risk and 35% moderate risk.

Figure 1 depicts the distribution of new confirmed cases of COVID-19 within the 14 days after the elections for both cohorts. A total of 37 cases (0.2%) were confirmed in the members of the polling station and 43 (0.23%) in the control group (p-value 0.576). Conversely, members of polling stations presented fewer cases within the 3 days after the elections: 3 (0.02%) versus 13 (0.07%) in the control group, p-value <0.05. No statistical differences were found between 3 and 14 days after the elections: 34 (0.19%) cases in the polling station members group and 30 (0.16%) in the control group, p-value=0.707.

**Figure 1.**
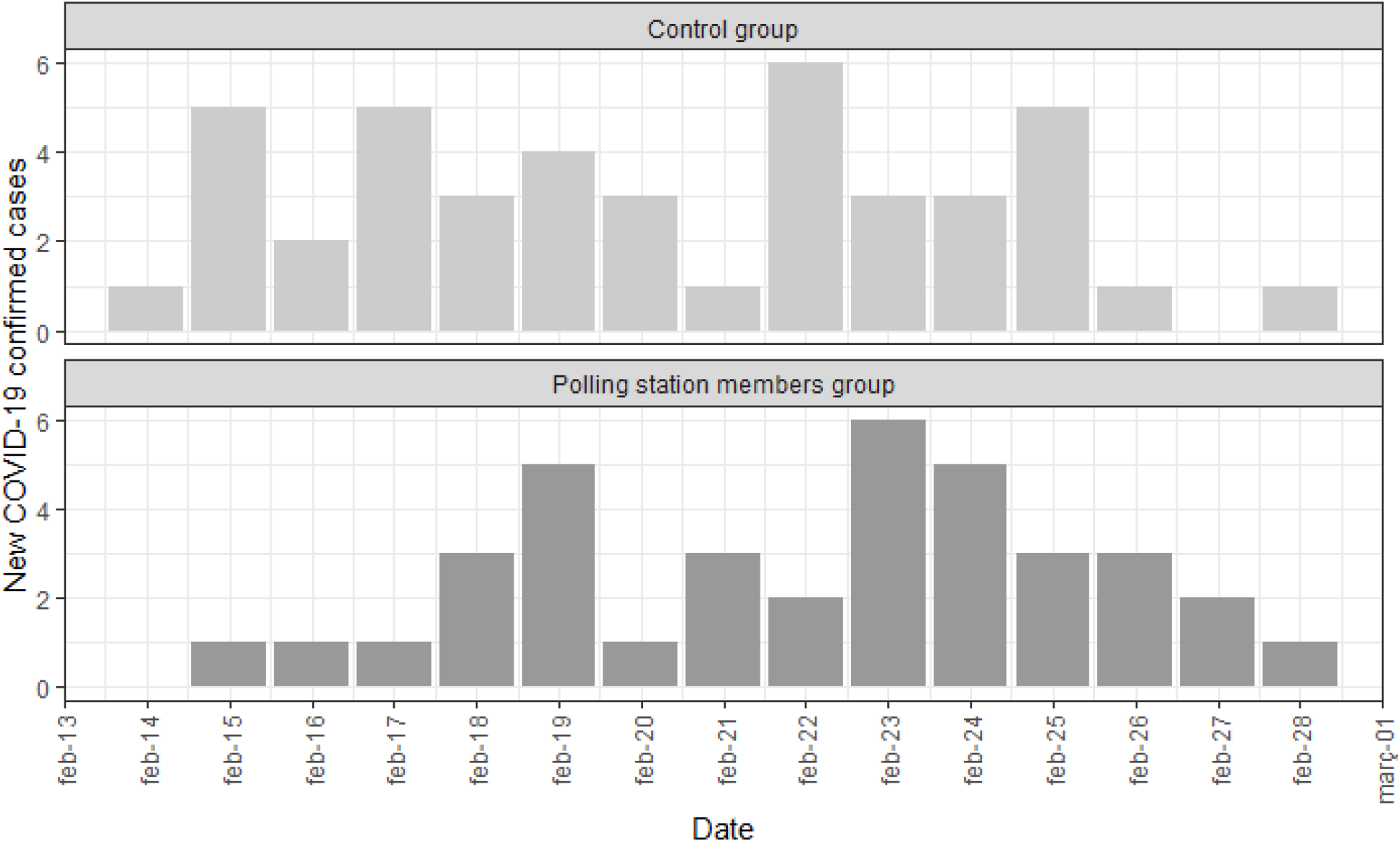
Distribution of new COVID-19 confirmed cases from 14th to 28th February 2021 for both cohorts.

## Discussion

We hereby report on the possible risk of SARS-CoV-2 transmission for the members of polling stations during the voting day in Catalonia. 14-day cumulative incidence in Catalonia that day was 340.74 cases per 100,000 inhabitants [6]. Even so, our analysis didn’t find any difference on the number of COVID-19 confirmed cases after the elections between a sample of members of polling stations and a control group. Our data suggest that members of polling stations were not at increased risk of infection. This finding is important for future decisions of holding elections elsewhere, as in some other places a percentage of polling stations were closed due to lack of available poll members as they feared to be infected by SARS-CoV-2 [7].

In addition, we also observed that polling stations members had less cases within the 3 days after the voting day than the control group. That could be related to the fact that the Catalan Government offered a LFT to all members of polling stations one or two days before the elections and those who tested positive were excluded from the polling station committees. This strategy could lead to a less infected population and thus reducing the risks during the voting day. This could be a measure to consider in further electoral procedures worldwide.

Our study has limitations. We do not have access to the entire census of people who were part of the polling stations during the Catalan elections and we ignore if there may be any bias in our sample. However we analysed data of more than 65% of polling stations and the control group was paired by place of residence. In addition we do not know the place where people in our study were infected.

Considering the importance of elections and the damages and dangers in democratic terms that a cancellation or suspension could have, our study suggests that if the appropriate measures are implemented, there is no greater risk of infection for the members of the polling stations, even in a situation of high community transmission of SARS-CoV-2.

## Data Availability

The data underlying this article will be shared on reasonable request to the corresponding author.

## Availability of data and materials

The data underlying this article will be shared on reasonable request to the corresponding author.

## Key-points

- During the COVID-19 pandemic some elections have been cancelled worldwide.
- We found non-differential risk of SARS-CoV-2 infection for polling station members during on Catalan elections.
- Offering a lateral flow to all members of polling stations before the voting day could be an action to consider to reduce the risk of infection.

